# Equations for Estimating Basal and Resting Metabolic Rates: A Scoping Review Protocol

**DOI:** 10.1101/2024.10.03.24314859

**Authors:** Veronika Pourová, Alena Langaufová, Abanoub Riad

**Affiliations:** Department of Public Health, Faculty of Medicine, Masaryk University, Brno, Czech Republic; Department of Health Sciences, Faculty of Medicine, Masaryk University, Brno, Czech Republic; Masaryk Centre for Global Health (MCGH), Department of Public Health, Faculty of Medicine, Masaryk University, Brno, Czech Republic

**Keywords:** Basal Metabolic Rate, Energy Expenditure, Evidence-Based Healthcare, Indirect Calorimetry, Resting Metabolic Rate

## Abstract

**Objective:** This scoping review aims to map the existing equations used for estimating basal and resting metabolic rates and to specify their applicability to various population groups, particularly in cases where they demonstrate higher accuracy based on age, ethnicity, and body mass index (BMI).

**Methods:** The proposed scoping review will be conducted in accordance with the JBI methodology for scoping reviews and reported following the Preferred Reporting Items for Systematic Reviews and Meta-Analyses for Scoping Reviews (PRISMA-ScR). A comprehensive search strategy has been developed to identify both published and unpublished studies. The literature search will be conducted in the following databases: MEDLINE (Ovid), EMBASE (Ovid), SportDiscus (EBSCO), and ClinicalTrials.gov. Abstracts will be screened against the inclusion criteria, and the full text of all potential studies will be reviewed by two independent researchers. Data will be extracted using a pre-designed data extraction sheet and presented in tabular form and as a narrative summary.

**Results:** The findings of this review will be disseminated through publication in a peer-reviewed journal and presented as an abstract at specialty conferences.

**Conclusion:** To the best of the authors’ knowledge, this review will be the first to comprehensively map all existing equations for both basal and resting metabolic rates. It is also a pioneering effort in synthesising evidence on the accuracy of these equations.

**Open Science Framework (OSF) Registration:** This project has been registered at OSF since October 3^rd^, 2024. https://doi.org/10.17605/OSF.IO/UD4WB

## Introduction

### Rationale

The treatment of obesity is multifaceted and requires the involvement of a multidisciplinary team, including dietitians, physiotherapists, physicians, and psychologists. Obese patients often experience a history of repeated cycles of weight loss and regain, making the accurate assessment of their energy needs crucial for developing appropriate dietary plans.^1^ Understanding these energy requirements is essential for setting an adequate energy intake that preserves muscle mass, supports overall tissue function, and reduces excess body fat.^1,2^

One of the most accurate methods for measuring resting energy expenditure (REE) is indirect calorimetry, which provides a direct measurement of a patient’s metabolic rate.^3^ However, this method requires significant technical and clinical expertise, and its application is relatively time-consuming and costly.^3,4^ In 2011, it was accessible to only about 18% of nutrition professionals in the UK.^5^ As such, its routine use in clinical practice is limited by resource constraints.^3–5^

In clinical settings where indirect calorimetry is unavailable, predictive equations are commonly employed to estimate REE and inform dietary planning. These equations typically consider variables such as body weight, height, age, and sex, with some incorporating additional factors such as fat-free mass and disease-specific parameters. For these equations to be deemed accurate, they should exhibit an error rate of less than 10% when compared to indirect calorimetry. However, the current literature indicates that even the most accurate predictive equations achieve this level of precision in only 73% of patients within specific body mass index (BMI) categories.^6–8^

Selecting the appropriate equation for a given patient population is critical to ensuring accurate estimation of energy needs when indirect calorimetry is not feasible. Therefore, a systematic and comprehensive review of existing predictive equations, including their accuracy and applicability to specific patient subgroups, is urgently needed to guide clinical decision-making in the management of obesity.

### Review Question(s)

This scoping review aims to explore the current landscape of equations used to estimate basal and resting metabolic rates (BMR and RMR) and their accuracy across different population groups. Therefore, it was designed to answer the following questions:

I. What are the existing equations used to estimate BMR and RMR?
II. Which population groups have been studied in the development of equations for estimating BMR and RMR?
III. How do these equations differ in terms of accuracy across specific population groups?

## Inclusion Criteria

### Participants

This review will include studies involving individuals from all demographic groups, with no exclusions based on age, gender, ethnicity, medical history, or BMI, thereby ensuring comprehensive coverage of diverse populations.

### Concept

This review will explore the various equations used to estimate basal and BMR and RMR and how they compare to indirect calorimetry. The focus will be on identifying, mapping, and synthesising evidence related to the accuracy and applicability of these equations in estimating resting energy expenditure (REE) across diverse populations.

### Context

This review will consider studies conducted in any geographic location and in various settings, including clinical, non-clinical, and research environments. It will account for factors such as cultural, racial, and gender influences that may impact metabolic rate estimates, aiming to provide a comprehensive understanding of how these equations perform across different population groups and contexts.

### Types of Sources

This review will include all analytical observational studies, such as cross-sectional, case-control, and cohort studies, that have utilised equations for measuring BMR and RMR. Eligible studies should compare the equations with indirect calorimetry.

## Methods

The proposed scoping review will be conducted according to the Joanna Briggs Institute (JBI) methodology for scoping reviews and will be reported as per the Preferred Reporting Items for Systematic Reviews and Meta-analyses for Scoping Reviews (PRISMA-ScR).^9,10^

### Search Strategy

A comprehensive three-phase search strategy was developed to locate both published and unpublished studies. In the first phase, an initial limited search of MEDLINE (Ovid) was conducted to identify relevant articles. In the second phase, a detailed literature search was carried out across multiple databases, including MEDLINE (Ovid), EMBASE (Ovid), SportDiscus (EBSCO), and ClinicalTrials.gov, using all identified keywords and index terms.

To cover grey literature, sources such as SportDiscus and ClinicalTrials.gov were included. The search focused on identifying all studies related to “resting energy expenditure” and “predictive equations.” No language or publication restrictions were applied to ensure a comprehensive search.

In the final phase, citation tracking will be conducted, where the reference lists of all studies meeting the inclusion criteria will be checked for additional relevant studies.

### Selection Process

Two independent reviewers will screen all retrieved articles in accordance with the inclusion criteria. In the initial screening phase, articles will be assessed based on their titles and abstracts. In the second phase, full-text screening will be conducted to determine the final inclusion of studies in the scoping review. Any disagreements between the two reviewers during the screening process will be resolved through consultation with a third reviewer. The study selection process will be illustrated using a PRISMA flow diagram, detailing each stage of the review.

### Data Extraction

Data will be extracted from all included studies using a structured data extraction sheet designed specifically for this review. The extracted data will include study characteristics (e.g., author, year of publication, study design, and sample size), details of the predictive equations for BMR and RMR, comparison with indirect calorimetry, and reported outcomes related to accuracy and applicability.

Two independent reviewers will perform the data extraction to ensure consistency and accuracy, resolving any discrepancies through discussion or by consulting a third reviewer if necessary.

### Data Analysis & Presentation

Data from the included studies will be synthesised qualitatively and will be presented in tabular form, accompanied by a narrative summary to highlight the accuracy and applicability of the various equations for estimating BMR and RMR. Graphical representations may be used to visually summarise trends and differences across population groups.

## Supporting information

Appendix I: Search Strategy

Appendix II: PRISMA-P Checklist

## Data Availability

All data produced in the present study are available upon reasonable request to the authors.

## Acknowledgements

The authors would like to thank Masaryk University Campus Library (KUK) for providing access to the necessary databases essential for conducting this scoping review.

## Appendices

*Appendix I: Search Strategy Appendix*

*Appendix II: PRISMA-P Checklist*

