## Appendix I: Search Strategy for "Equations for Estimating Basal and Resting Metabolic Rates: A Scoping Review Protocol"

**Ovid MEDLINE(R) ALL 1946 to August 05, 2024**

**The search was conducted on 6^th^ August 2024 at 9:00 am (CET).**

|  | Searches | Results |
| --- | --- | --- |
| 1 | energy metabolism/ or basal metabolism/ | 98371 |
| 2 | ((metabol* or expenditure?) adj1 energ*).kf,kw,tw. | 85593 |
| 3 | ((resting or basal or basic) adj1 metabol*).kf,kw,tw. | 10651 |
| 4 | (bioenergetic* or BMR or RMR or basal oxygen consumption?).kf,kw,tw. | 19663 |
| 5 | or/1-4 | 163085 |
| 6 | "Predictive Value of Tests"/ | 227092 |
| 7 | (predict* adj3 (equation* or formula? or calculation? or model*)).kf,kw,tw. | 225913 |
| 8 | (predict* adj1 (test? or value?)).kf,kw,tw. | 166522 |
| 9 | or/6-8 | 557677 |
| 10 | 5 and 9 | 3514 |

**Embase 1974 to 2024 August 05**

**The search was conducted on 6^th^ August 2024 at 10:00 am (CET).**

| **#** | **search string** | **# of results** |
| --- | --- | --- |
| 1 | basal metabolic rate/ or resting metabolic rate/ or resting energy expenditure/ | 13373 |
| 2 | ((metabol* or expenditure?) adj1 energ*).kf,kw,tw. | 106014 |
| 3 | ((resting or basal or basic) adj1 metabol*).kf,kw,tw. | 12666 |
| 4 | (bioenergetic* or BMR or RMR or basal oxygen consumption?).kf,kw,tw. | 25999 |
| 5 | or/1-4 | 138219 |
| 6 | Predictive Value/ | 271829 |
| 7 | (predict* adj3 (equation* or formula? or calculation? or model*)).kf,kw,tw. | 285191 |
| 8 | (predict* adj1 (test? or value?)).kf,kw,tw. | 248102 |
| 9 | or/6-8 | 642233 |
| 10 | 5 and 9 | 3247 |

**SPORTDiscus with Full Text**

**The search was conducted on 6^th^ August 2024 at 10:35 am (CET).**

| **#** | **search string** | **# of results** |
| --- | --- | --- |
| **1** | (MH"Basal Metabolism" OR TI (((metabol* OR expenditure#) N2 energ*) OR ((resting OR basal OR basic) N2 metabol*) OR (bioenergetic* OR BMR OR RMR OR “basal oxygen consumption#”)) OR AB (((metabol* OR expenditure#) N2 energ*) OR ((resting OR basal OR basic) N2 metabol*) OR (bioenergetic* OR BMR OR RMR OR “basal oxygen consumption#”))) AND (TI ((predict* N4 (equation* OR formula# OR calculation# OR model*)) OR (predict* N2 (test# or value#)) OR AB ((predict* N4 (equation* OR formula# OR calculation# OR model*)) OR (predict* N2 (test# or value#)))) | 332 |

**ClinicalTrials.gov**

**The search was conducted on 6th August 2021 at 11:00–11:30 am (CET).**

| **#** | **search string** | **# of results** |
| --- | --- | --- |
| **1** | “basal metabolism” OR “basic metabolism” OR “resting metabolism” OR “basic energy expenditure” OR “resting energy expenditure” – **in Field Condition or disease** |  |
| **2** | equation OR formula OR model OR calculation OR equations OR models OR formulas OR calculations-**in Field Other terms** |  |
| **3** | 1 AND 2 | 624 |
